# Easy-BILAG: a new tool for simplified recording of SLE disease activity using BILAG-2004 index

**DOI:** 10.1101/2021.07.30.21261385

**Authors:** Lucy M Carter, Caroline Gordon, Chee Seng Yee, Ian Bruce, David Isenberg, Sarah Skeoch, Edward M Vital

## Abstract

**Objective:** BILAG-2004 index is a comprehensive disease activity instrument for SLE but administrative burden and frequency of errors limits its use in routine practice. We aimed to develop a tool for more accurate, time-efficient scoring of BILAG-2004 index with full fidelity to the existing instrument.

**Methods:** Frequency of BILAG-2004 items was collated from a BILAG-biologics registry (BILAG-BR) dataset. Easy-BILAG prototypes were drafted to address known issues affecting speed and accuracy. After expert-verification, accuracy and usability of the finalised Easy-BILAG was validated against standard format BILAG-2004 index in a workbook exercise of 10 case vignettes. 33 professionals with a range of expertise from 14 UK centres completed the validation exercise.

**Results:** Easy-BILAG incorporates all items present in ≥5% BILAG-BR records, plus full constitutional and renal domains into a rapid single-page assessment. An embedded glossary and colour-coding assists scoring each domain. A second page captures rarer manifestations when needed. In the validation exercise, Easy-BILAG yielded higher median scoring accuracy (96.7%) than standard BILAG-2004 documentation (87.8%, p=0.001), with better inter-rater agreement. Easy-BILAG was completed faster (59.5min) than the standard format (80.0min, p=0.04) for 10 cases. An advantage in accuracy was observed with Easy-BILAG use among general hospital rheumatologists (91.3 vs 75.0, p=0.02), leading to equivalent accuracy as tertiary centre rheumatologists. Clinicians rated Easy-BILAG as intuitive, convenient, and well adapted for routine practice.

**Conclusion:** Easy-BILAG facilitates more rapid and accurate scoring of BILAG-2004 across all clinical settings which could improve patient care and biologics prescribing. Easy-BILAG should be adopted wherever BILAG-2004 assessment is required.

## Introduction

Disease activity measurements in systemic lupus erythematosus (SLE) are necessary for optimal patient care. They are central to clinical guidelines(1, 2) and treat-to-target approaches, which have been shown to improve outcomes in SLE(3, 4), rely on specifically defining and measuring low disease activity and remission(5). Furthermore, national commissioning policies for biologic agents also increasingly stipulate measured baseline and response disease activity criteria (2, 6).

Recording complex multisystem manifestations longitudinally is a significant challenge in SLE. As a result, formalised disease measures have been less readily embedded in routine care(7) compared with other rheumatic diseases such as rheumatoid arthritis. Composite disease activity instruments, including the SLEDAI, ECLAM and the British Isles Lupus Assessment Group (BILAG) -2004 index, all have proven validity and reliability(2, 8, 9). The BILAG-2004 index is the most comprehensive available instrument. It replaced the original ‘classic’ BILAG index (10) and the current version includes numerical scoring and updates to haematology items (11, 12). BILAG-2004 index comprises 97 discrete clinical manifestations of SLE across 9 organ domains. The activity in each domain is graded separately from A, highly active and likely to necessitate escalation in therapy, to E, no current or previous disease activity (13). BILAG-2004 captures several important disease features such as haemolysis, lymphadenopathy, polyneuropathy, interstitial pneumonitis, gastrointestinal and significant although rare ophthalmogical items which do not feature in SLEDAI. Additionally, all organ domains carry potentially equal weighting. Unlike SLEDAI, BILAG-2004 index differentiates between disease features which have partially, but not completely improved, those which have not changed and those which are worse. It is thus more sensitive than SLEDAI to changes in disease activity over time and better detects partial response to therapy and the exacerbation of already active disease features(14, 15). These differences may be particularly important in research studies.

The current BILAG-2004 documentation relies on an index case report form, a detailed glossary of clinical items and a separate scoring algorithm for each of the 9 organ domains. Formal training is recommended. Therefore, despite its advantages, BILAG-2004 index may be difficult or time-consuming to complete during routine clinic visits, particularly for those not familiar with the glossary and the layout of the case report form and scoring document, even though most patients have relatively few abnormal items present at any single visit.

The Easy-BILAG project aimed to develop and validate a simplified tool to record and score the current published version of the BILAG-2004 index(11) more rapidly and accurately for use in routine clinical care.

## Methods

### Development process

Easy-BILAG was registered as a multi-centre quality improvement initiative with Quality Assurance and Governance department of Leeds Teaching Hospitals NHS Trust and at the relevant governance departments at individual participating Trusts. No real-patient was used in Easy-BILAG validation material and specific research ethics approval was therefore not obtained. BILAG-BR is an ongoing prospective study with research ethics approval from NRES Committee North West–Greater Manchester West (REC: 09/H1014/64) and Health Research Authority approved on 9 November 2009 (IRAS ref. 24407).

Reasons underlying inaccuracy or difficulty completing BILAG-2004 assessment were discussed in meetings of the BILAG group of expert clinicians, experienced in delivering BILAG-2004 training and adjudicating clinical trials. The frequency of BILAG-2004 clinical items was evaluated in an active SLE cohort. Based on these insights, a series of Easy-BILAG prototypes were developed to address the key problems identified. The finalised Easy-BILAG was validated in a workbook exercise. Minor changes to wording in the constitutional and renal domains were made after validation as directed by feedback from participants. A separate self-adjudication check list was also added, to assist with use in a clinical trial setting, after the validation.

### Determining the frequency of BILAG-2004 SLE manifestations

Pseudonymised BILAG-2004 disease activity scores, from individual SLE patients enrolled in the UK BILAG-Biologics Registry (BILAG-BR) between March 2010 and November 2019, were available for evaluation. The majority of enrolled patients had moderate-severe SLE disease activity and were commencing biologic therapy (16). The frequency with which each of the 97 BILAG-2004 clinical items was recorded as ‘new’, ‘improving’, ‘same’ or ‘worse’, was quantified. Quantitative items (i.e. full blood count, creatinine, eGFR, proteinuria and blood pressure) were recorded as raw numerical values in this dataset, independent of attribution to SLE disease activity and these were therefore were excluded from this analysis.

### Validation exercise

Consultant rheumatologists including members of BILAG as well as those without sub-speciality interests in SLE, Rheumatology specialty trainees and experienced lupus specialist nurses from 14 UK centres were invited by BILAG members to complete a timed validation workbook of 10 short case vignettes. Workbooks were distributed to participating centres by mail. Each participant was provided with a workbook which randomly assigned them to Easy-BILAG or standard format. Each clinician thereby scored BILAG-2004 disease activity for the same 10 cases using either Easy-BILAG or standard format BILAG-2004 index form, glossary and scoring algorithm (11). No prior BILAG-2004 training or experience was prerequisite. The validation workbook was designed to test clinicians in scoring both frequent and uncommon SLE manifestations, and test longitudinal scoring of items in flare and remission. All workbooks contained an introductory overview and detailed instructions on how to use BILAG-2004 index. Individuals self-reported time taken to score the 10-cases. They were also asked to report their level of prior BILAG-2004 experience, job role and whether they worked within a general or tertiary hospital setting. Overall perception and usability of the scoring format was assessed by 4 Likert-item survey questions. Clinicians returned their workbooks in hard copy to the central study team and could opt to submit anonymously. Item and domain level accuracy was calculated against a model answer scheme verified in advance by original BILAG-2004 authors.

### Statistical analysis

After demonstrating non-normality, variables were compared between two groups in SPSS using Wilcoxon Rank-Sum Test and between multiple groups by Kruskall Wallis followed by relevant pairwise comparisons. Inter-rater agreement for all domain scores was evaluated according to BILAG-2004 format by Fleiss’ kappa coefficient using R Studio *irr* package (17) and the result categorized as previously described (18). Statistical significance was considered p < 0.05 on one-tailed testing.

## Results

### Key problems identified in BILAG-2004 scoring

Consultation among BILAG-2004 lead authors and expert clinicians identified features of the current published recording format which pose barriers to its accurate and/or time-efficient use in routine practice, many of which are also relevant to clinical trials. Strategies to resolve priority issues were developed. These are collectively summarized in Supplementary Table 1.

### Item-level frequency of BILAG-2004 SLE manifestations in a UK biologics registry

Item level data from 2395 BILAG-BR disease activity records revealed that the 6 most frequent items were each present in more than 20% of records, namely: mild arthritis (72%), mild skin eruption (47%), moderate arthritis (38%), mild mucosal ulceration (34%), mild alopecia (34%) and pleurisy / pericarditis (22%). Twenty-two discrete items were present in 5% or more of cases and no Ophthalmic or Gastrointestinal domain items were among these. Twenty-five items were active in less than 1% of BILAG-BR records (Supplementary Fig.1).

### The Easy-BILAG template

Easy-BILAG was designed to enhance the visibility of the most frequently scored BILAG-2004 items identified by a BILAG-BR dataset (Fig.1). A single page Easy-BILAG now captures full disease activity for 68% of biologics-treated patients in BILAG-BR. The constitutional and renal domains are scored in full on page 1 and all other clinical items recorded as active in >5% of BILAG-BR records are also captured on the first page (Fig.1). Less frequent manifestations are scored, only when necessary, on a second page (Supplementary Fig.2) as prompted by screening questions on page 1 (Fig.2A). The design incorporates an abridged glossary definition immediately adjacent to clinical each item to facilitate closer adherence to glossary criteria. A colour-blindness compatible, colour-coding system directs clinicians instantly to the overall A-E score for each domain, so complete scores are readily derived at the time of assessment (Fig.2A-C). Constitutional and renal domains follow a similar principle, but, because a combination of features is needed to compute the domain score, we have adopted an arbitrary points system to facilitate scoring with full fidelity to original BILAG-2004 algorithms (Fig.2C). On completion of Easy-BILAG, an entirely optional self-adjudication checklist, ensuring compliance with core BILAG-2004 scoring rules such as attribution to SLE, improvement criteria and trickle-down rule is available and might assist particularly in a clinical trial setting (Supplementary Fig.3)

**Figure 1.**
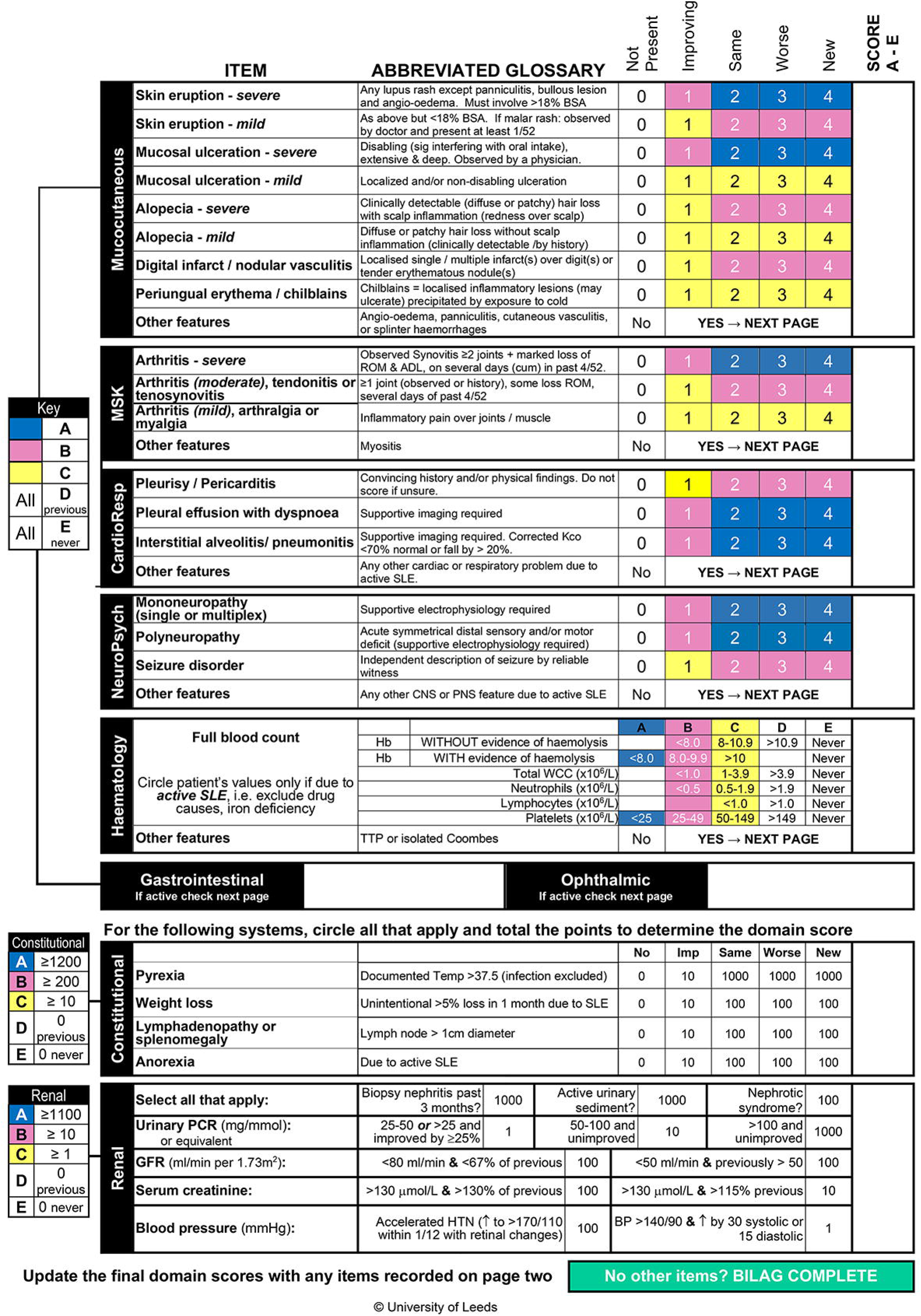
Easy-BILAG page 1 in full demonstrates structure of the scoring template for the most frequently encountered SLE features in addition to the constitutional and renal domains in full. Items are organised within tables by organ domain. Each is anchored to its relevant colour coded key to the left. Scoring of items from ‘not present’ (0) to ‘new’ (4) is colour coded and translates to overall organ domain score from A (blue), B (pink), C (yellow) to D or E (white). Free text space is included to the right of each domain for the clinician to assign the final domain score from A-E. An abbreviated glossary definition is included immediately adjacent to each item. Gastrointestinal and Ophthalmic domains prompt clinicians to consult page 2 of Easy-BILAG (See Supplementary Figure 2). Constitutional and Renal domains derive domains scores using a tally of arbitrary points assigned to each item. A separate key anchored to both domains directs overall scoring according to points tally. If no items on page 2 required to be scored the BILAG assessment is then complete. *MSK – musculoskeletal; CardioResp – cardiorespiratory; NeuroPsych – neuropsychiatric*.

**Figure 2.**
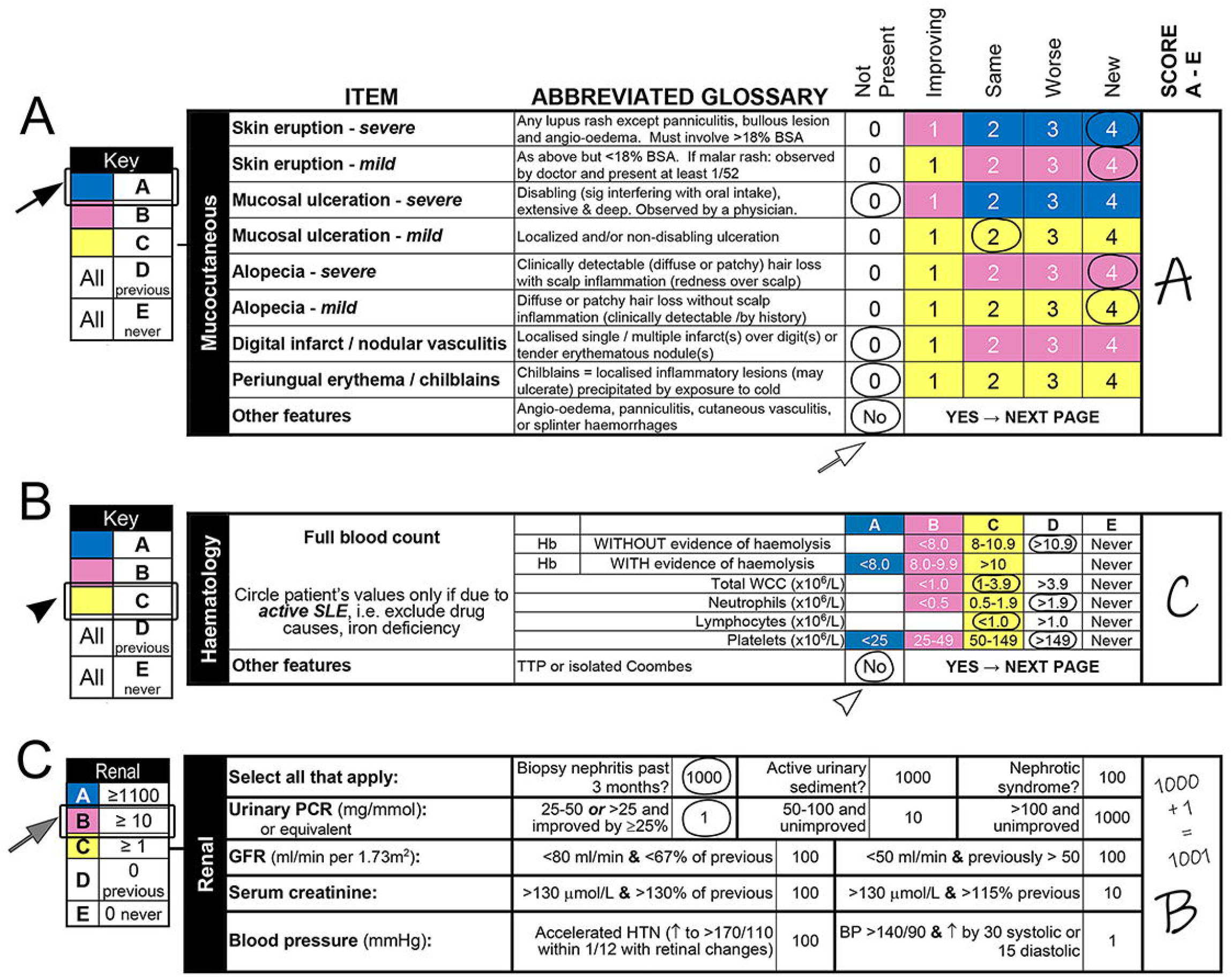
Annotated sections of Easy-BILAG illustrate method for deriving A-E scores for example domains. Standard BILAG-2004 principles apply throughout. Mucocutaneous scoring (A) requires clinician to circle each listed item as ‘not present’ (0), ‘improving’ (1), ‘same’ (2), ‘worse’ (3) or ‘new’ (4) and indicate whether any ‘Other features’ recorded on Easy-BILAG page 2 are present. Colour-coded key (left) directs to the overall domain score, in order of disease activity from A (blue; highly active); B (pink; moderately active); C (yellow; mild stable disease) to D (white; no disease activity, but previously present) or E (white; no disease activity and never previously active). The highest tariff item triggers the final domain score. Example shows ‘Skin eruption – severe’ recorded as ‘new’ (blue) triggering overall mucocutaneous domain score A (filled arrow) since no ‘Other features’ are present (open arrow). ‘A’ is then noted in the free text space (right). Haematological domain (B) requires clinicians to circle the range in which each full blood count differential is located and indicate whether ‘Other features’ of haematological SLE are present. Haemoglobin is scored separately according to whether haemolysis is present or absent. The severity of anaemia / cytopaenia translates to overall domain score based on the same colour-coded key (left) used elsewhere. Example shows ‘total white cell count of 1-3.9 ×10^9^/L’ and ‘lymphocyte count < 1.0 ×10^9^/L’ (yellow) with no ‘Other features’ present (open arrowhead) triggering overall haematological domain score C (filled arrowhead) which is noted in the free text space (right). Renal domain (C) requires clinicians to circle all features of renal disease which are present. Items are assigned arbitrary points (1, 10, 100 or 1000). The total points from items recorded translates to overall domain score based on a separate renal domain key anchored to the left. Example records ‘biopsy confirmed nephritis’ and ‘urine protein:creatinine ratio 25-50’ as present, totalling 1001 arbitrary points and triggering overall renal domain score B (grey arrow) which is noted in the free text space.

### Easy-BILAG facilitates superior BILAG-2004 scoring accuracy and speed

Accuracy of Easy-BILAG scoring was tested against standard format BILAG-2004 index in a validation workbook of 10 case vignettes completed by rheumatology professionals (n = 33) in 14 centres around the Great Britain and Northern Ireland. Forty-five percent of participants in the validation exercise were consultant rheumatologists or clinical academics and 45% were speciality trainees. Overall, 42.4% reported their use of BILAG-2004 index in their current practice as infrequent or rare. Further characteristics of professionals participating are shown in Supplementary Table 2.

Accuracy of scoring (% accuracy; *median* (*Quartile 1, Quartile 3*)) against expert verified model answers was significantly higher across all domains with use of Easy-BILAG (n = 16; 96.7 (94.4, 97.5), Mann-Whitney *U* = 53.0, p = 0.001; Fig.3A) as compared with the standard BILAG-2004 format (n = 17; 87.8 (80.0, 94.4); Fig.3A). Since assessment of active rather than quiescent disease is more prone to error, rating of domains requiring grade A to C scores was examined separately. Rating of active domains retained high levels of accuracy with use of Easy-BILAG (94.6 (89.1, 97.2); Fig.3B) which was significantly above that observed using standard format BILAG-2004 (80.4 (65.2, 90.2), *U* = 48.0, p = 0.001; Fig.3B). Self-reported completion time for the 10-case workbook (minutes; *median* (*Q*_*1*_, *Q*_*3*_)) was significantly shorter among clinicians using Easy-BILAG (n = 16; 59.5 min (53.2, 86.3); Fig.3C) than standard format (n = 17; 80.0 min (61.0, 104.0), *U* = 87.0, p =0.04; Fig.3C).

**Figure 3.**
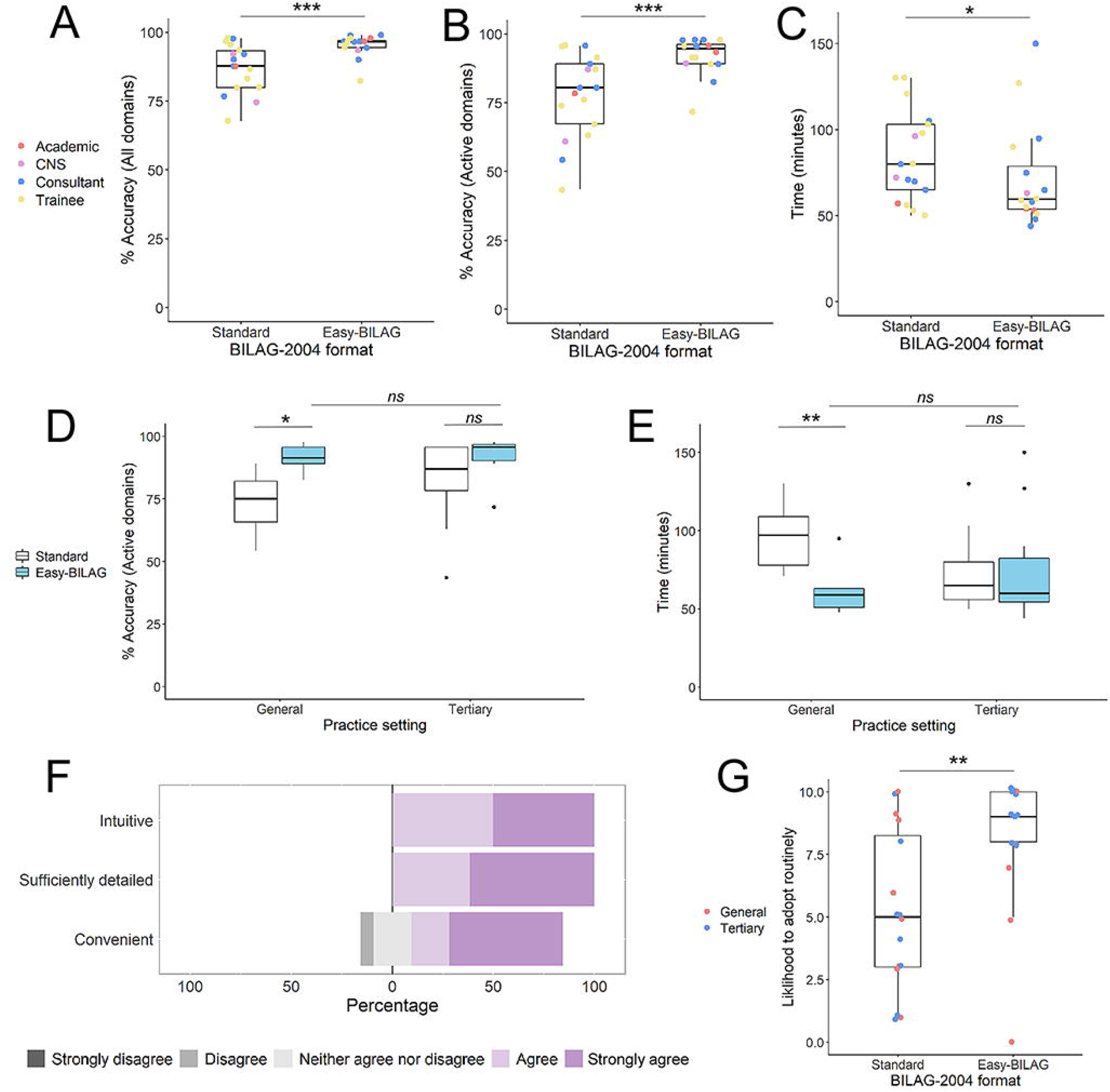
Boxplots plots (A-C) show performance of Easy-BILAG against standard format BILAG-2004 in self-timed validation workbook exercise for all professional categories (overlay jitter points). Scoring accuracy (%) against model workbook answers for all organ domains (A) and active organ domains (B) was significantly higher with Easy-BILAG. Time taken to complete workbook exercise (C) was significantly shorter with Easy-BILAG. Boxplots (D-E) show Easy-BILAG (blue) versus standard format BILAG-2004 (white) across general and tertiary practice. Scoring accuracy (D) was significantly higher and time taken (E) reduced among general hospital rheumatologists using Easy-BILAG and equivalent to tertiary rheumatologists. Stacked horizontal bar chart (F) shows Easy-BILAG rated favourably for intuitiveness, detail and convenience on 5-point Likert-scale. Boxplot (G) shows clinicians in tertiary and general rheumatology (overlay jitter points) reported significantly higher likelihood to adopt instrument in routine practice was significantly higher by 10-point visual analogue scale (10 highly likely, 0 highly unlikely) for Easy-BILAG was significantly by clinicians across general and tertiary practice (overlay jitter). *** p ≤ 0.001; ** p ≤ 0.01; p ≤ 0.05; ns – non significant, p > 0.05

### Easy-BILAG matches general hospital and tertiary centre clinicians for accuracy and speed

The performance of Easy-BILAG outside of centralised, subspeciality and research-orientated tertiary centres was evaluated. Accuracy in grading active domains (% accuracy; *median* (*Q*_*1*_, *Q*_*3*_)) was high among tertiary centre clinicians using standard format BILAG-2004 (n = 9; 86.9 (70.7, 95.7) Fig. 3D) but still trended to higher accuracy among those using Easy-BILAG (n =11; 95.7 (90.2, 96.7), p = 0.06 Fig.3D). In contrast, general hospital clinicians using standard format BILAG-2004 returned significantly lower accuracy (n = 8; 75.0 (64.1, 83.7)) than those testing Easy-BILAG (n = 5; 91.3 (90.2,96.7), p = 0.02; Fig.3D) while the latter achieved accuracy comparable with tertiary centre colleagues (p = 0.70). Among general hospital clinicians, workbook completion was significantly faster (time in minutes; *median* (*Q*_*1*_, *Q*_*3*_)) with Easy-BILAG (n = 5; 59.0 (51.0, 63.0), Fig.3E) than with standard format (n = 8; 97.0 (76.0, 113.0), p = 0.01; Fig.3E). Easy-BILAG achieved similarly high and matched levels of accuracy between clinicians with and without prior BILAG-2004 training, and between clinicians who regularly or infrequently use BILAG-2004 in their existing practice (Supplementary Figure 4).

### Easy-BILAG improved inter-rater agreement on disease activity

Overall inter-rater agreement in workbook disease activity grading using standard BILAG-2004 format was classified as good, though levels of agreement in assigning active grade B disease were lower (Table 1). Clinicians testing Easy-BILAG demonstrated higher levels of inter-rater agreement overall and across each level of active disease graded A-C where agreement was classified as very good (Table 1).

**Table 1.**
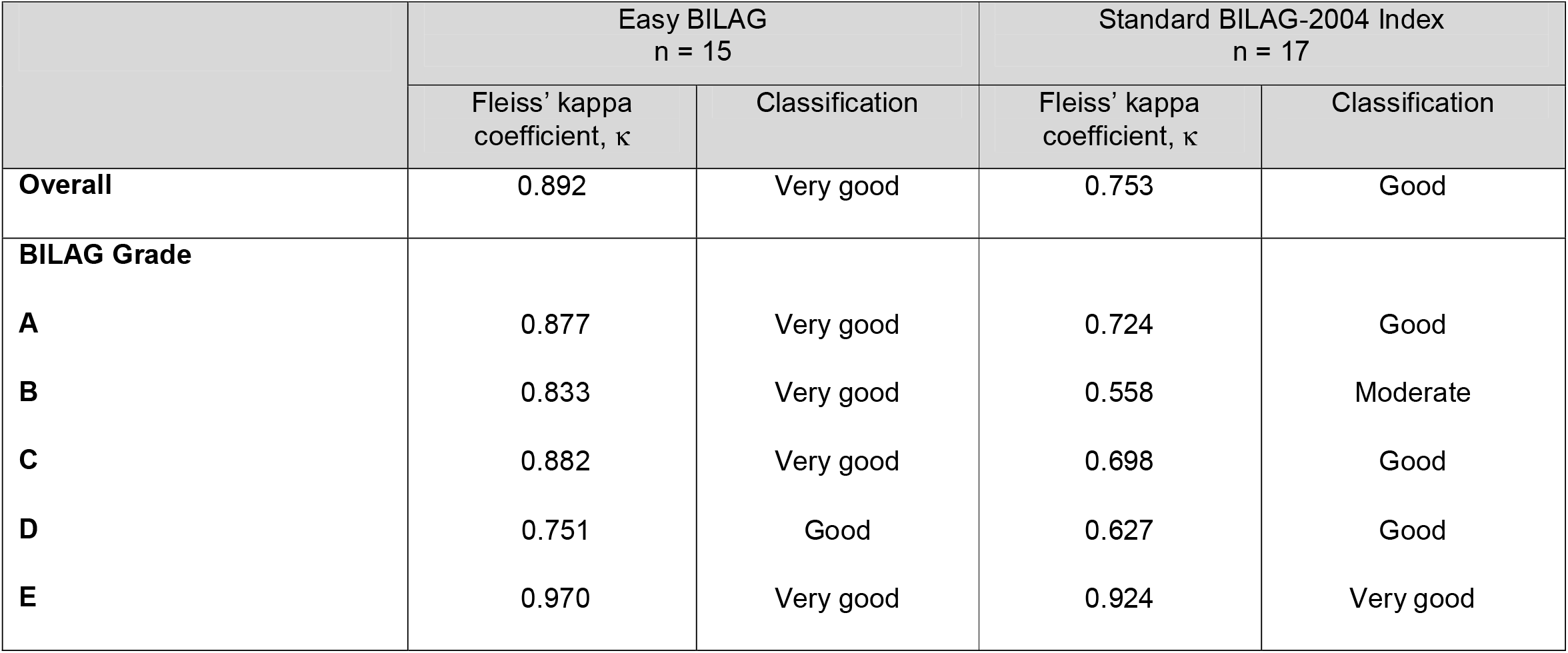
Inter-rater agreement in BILAG-2004 domain scoring by validation workbook format.

|  |  |  |  |  |
| --- | --- | --- | --- | --- |
|  | Easy BILAG<br>n = 15 |  | Standard BILAG-2004 Index<br>n = 17 |  |
| | Fleiss' kappa<br>coefficient, $\kappa$ | Classification | Fleiss' kappa<br>coefficient, $\kappa$ | Classification |
| **Overall** | 0.892 | Very good | 0.753 | Good |
| **BILAG Grade** |  |  |  |  |
| **A** | 0.877 | Very good | 0.724 | Good |
| **B** | 0.833 | Very good | 0.558 | Moderate |
| **C** | 0.882 | Very good | 0.698 | Good |
| **D** | 0.751 | Good | 0.627 | Good |
| **E** | 0.970 | Very good | 0.924 | Very good |

### Easy-BILAG has high usability for routine practice

Clinician feedback was surveyed by Likert scale responses on completion of the validation exercise. All clinicians testing Easy-BILAG agreed that the format was intuitive and sufficiently detailed, while 12/16 (75.0%) agreed Easy-BILAG was a convenient format for disease activity recording (Fig.3F). By visual analogue scale (VAS 0 – highly unlikely; 10 – highly likely; median, (*Q*_*1*_, *Q*_*3*_)), clinicians from both tertiary and general hospitals testing Easy-BILAG reported significantly higher likelihood of adopting the tool in regular practice (n = 16, VAS: 8.5 (7.5, 10.0); Fig.3G) than those using standard format BILAG-2004 (n = 16 VAS: 5.0 (3.0, 8.5); U = 68.0, p = 0.01).

## Discussion

We report a novel tool for recording BILAG-2004 disease activity index which combines full fidelity to the current instrument (11) with a condensed, data driven, colour-coded design adapted to assist clinicians assess SLE in routine practice. In this validation exercise it yielded superior accuracy and time-efficiency over current standard format BILAG-2004 index using paper forms, with a particularly marked advantage among clinicians based outside of tertiary centres. It showed improved inter-rater reliability and perceived usability for routine practice among clinicians of varying levels of prior experience across different hospital settings. Thus Easy-BILAG offers a tool with which clinicians can confidently, accurately and time efficiently integrate BILAG-2004 index into routine practice.

Treat-to-target principles have demonstrated wide ranging benefits on patient outcomes across rheumatic diseases and it is clear that this approach reduces both flare rates and damage accrual in SLE(4). To target remission and low disease activity states, clinicians must be equipped with tools that allow them to measure disease activity accurately and consistently. BILAG-2004 index is a highly comprehensive disease activity instrument which assures clinicians that they have performed a thorough SLE assessment. It can therefore support and prompt clinical decision making, but its administrative burden is high and inadequately adapted to routine practice. The other major disease activity instrument, SLEDAI, is often felt to be the quicker and easier instrument (9) as it offers a limited item, fixed scoring format. It does however have different limitations, for example, some features such as autoimmune haemolysis are not scored at all, and due to the fixed points weighting, features such as lupus rash or thrombocytopaenia, however severe, can never in isolation translate to the high disease activity score. Further, arthritis always scores twice as highly as rash even if a mild arthritis and severe rash are present. Therefore SLEDAI does not always well align with the patient experience and physician’s intention to treat (15). By re-ordering items by their frequency and screening questions for rarer manifestations Easy-BILAG offers a balance between the simplicity and speed of the SLEDAI for common features while retaining the sensitivity to change and scope of BILAG-2004.

User feedback indicates Easy-BILAG had substantially greater appeal and usability for routine practice than standard format BILAG-2004 index. Influential to this we found Easy-BILAG permitted significant time saving for clinicians. Using Easy-BILAG, professionals with a range of prior BILAG-2004 experience completed 10 validation case assessments in an average time of just below 6 minutes per case. This time accounts for reading and evaluating the case material, and the cases were designed to cover both rare and common manifestations, real-world time required to document and derive a score for a familiar patient is likely to be very much less. Completing BILAG-2004 index accurately in standard published format requires consulting a clinical glossary and a separate scoring algorithm for each organ domain. Easy-BILAG makes use of colour coding and abridged glossary descriptions to reduce reliance on separate reference documents. Using data-driven design, the same BILAG-2004 assessment would require only a single page assessment for the majority of UK registry patients. Since its design was informed by item frequency in a biologics registry it is likely that far more routine outpatient visits would be less complex and quicker.

Easy-BILAG facilitated excellent scoring accuracy when tested against model case vignettes. Crucially, a similarly high level of accuracy was achieved by clinicians based in general hospital practice compared with those in tertiary centres where more subspecialist and research-focused activities using BILAG-2004 are typically concentrated. We also found that similar accuracy was maintained irrespective of prior training or current use of BILAG-2004 index. It is important to note that our validation exercise included a detailed overview and scoring instructions for BILAG-2004 index. Therefore Easy-BILAG is not a replacement for appropriate training, but our findings do suggest it is an accessible format for clinicians across all major areas of practice and can be more readily applied by those with less prior experience.

Easy BILAG not only showed higher scoring accuracy, but also substantially less variability. We found that in this group of real-world clinicians with a range of prior BILAG-2004 experience, Easy-BILAG achieved better inter-rater agreement across all disease activity grades than standard format scoring. Importantly variation was not simply a function of clinical role, practice setting or prior experience as Easy-BILAG appeared to show an advantage among all professional categories. In practice this has implications for continuity of care where a team of various clinicians may review or manage an individual during follow up and make treatment decisions such as whether to continue a biologic therapy. In the validation exercises, scoring with standard format appeared particularly vulnerable to inconsistencies assigning scores to grade B disease which has also been observed in defining moderate flares (19). Easy-BILAG was particularly beneficial in these cases and among these mixed-experience clinicians. Since grade B disease in two or more organ domains defines moderate disease activity in current EULAR, BSR and NHS England guidelines (1, 2, 6), this is a particularly decisive aspect of scoring where inconsistencies could conceivably introduce unwarranted variation in access to therapies. Improved recording accuracy might improve the identification of flares in clinical practice and the appropriate referral for new therapies.

Although our main goal in development of the Easy-BILAG was for use in routine clinical practice, our findings may have implications for clinical studies. Robust disease activity grading is central to trial outcome measures and the success or failure of new therapies in SLE. While detailed training in BILAG-2004 is essential for clinical investigators, we found that even among experienced lupus clinicians BILAG-2004 accuracy can be enhanced and variability reduced by formatting the assessment in a novel way. Investigators should be mindful of how data collection tools are presented to clinicians and the example of Easy-BILAG may be relevant to disease areas beyond SLE. We would also anticipate that integrating BILAG-2004 into more routine practice through use of Easy-BILAG could facilitate more robust data collection in clinical practice and better enable clinicians to take part in clinical trials and identify eligible patients.

The current work has some limitations. Firstly, due to constraints on space not all glossary items could be incorporated for rare items scored on Easy-BILAG page 2 and clinicians would still need to refer to core BILAG-2004 official glossary if assessing these features. Secondly as validation work was conducted on expert standardised training material, Easy-BILAG is yet to be evaluated in real world practice with real patients. Additionally, applying criteria for improvement and, worsening criteria as well as the ‘trickle down’ rule all still require appropriate understanding and training on BILAG-2004 index. An optional self-adjudication checklist at the end of Easy-BILAG has been devised as a prompt for these areas. Computer-assisted formats with inbuilt quality assurance on all aspects of BILAG-2004 can offer additional error checking which is particularly valuable in clinical trials (10).

In conclusion, Easy-BILAG will increase the use of BILAG-2004 in routine practice. This is anticipated to improve comprehensive and consistent assessment of patients, detection of disease activity, appropriate prescribing of immunosuppressants and biologics and judicious use of glucocorticoids. It is our recommended format for recording BILAG-2004 disease activity assessments in future clinical practice.

## Data Availability

No data are available.

## Acknowledgements

This research was supported by the United Kingdom National Institute for Health Research Leeds Biomedical Research Centre based at the Leeds Teaching Hospitals NHS Trust. The views expressed are those of the authors and not necessarily those of the NHS, the NIHR or the Department of Health.

We thank rheumatology clinicians from the following centres for their participation in the Easy-BILAG validation exercise: Leeds Teaching Hospitals NHS Trust, Sandwell and West Birmingham NHS Foundation Trust, Doncaster & Bassetlaw Teaching Hospitals NHS Foundation Trust, Royal National Hospital for Rheumatic Diseases, Oxford University Hospitals NHS Foundation Trust, Royal Berkshire NHS Foundation Trust, Frimley Health NHS Foundation Trust – Wexham Park, University College London Hospitals NHS Foundation Trust, University Hospitals Birmingham NHS Foundation Trust, Manchester University NHS Foundation Trust, East Lancashire Hospitals NHS Trust - Royal Blackburn Teaching Hospital, Belfast Health and Social Care Trust - Musgrave Park, Cambridge University Hospitals NHS Foundation Trust and Sheffield Teaching Hospitals NHS Foundation Trust.

Easy-BILAG is copyright of Edward Vital, University of Leeds. It is available for free use for routine clinical practice and academic studies, and for commercial purposes under license at https://licensing.leeds.ac.uk/products/healthcare-questionnaires

## Key Messages

- Easy-BILAG is a high-accuracy, time efficient tool for recording BILAG-2004 disease activity in SLE.
- It is the new recommended format for scoring BILAG-2004 index in clinical practice.
- Easy-BILAG and its training material is available free of charge for use in routine care at https://licensing.leeds.ac.uk/products/healthcare-questionnaires.

