## Supplementary Material for "Easy-BILAG: a new tool for simplified recording of SLE disease activity using BILAG-2004 index"

Contents:

1. Table S1: Key barriers to accurate routine use of BILAG-2004
2. Figure S1: Frequency of BILAG-2004 clinical items in a biologics registry (BILAG-BR) cohort BILAG-BR.
3. Figure S2: Easy-BILAG page 2 captures infrequent and rare BILAG-2004 clinical items.
4. Figure S3: Optional Easy-BILAG self-adjudication checklist.
5. Table S2: Characteristics of professionals participating in Easy-BILAG validation exercise.
6. Figure S4: Accuracy of Easy-BILAG by prior BILAG-2004 training and experience.

| Supplementary table S1. Expert -identified priority themes for improving routine usability of BILAG-2004 index | | | |
|---|---|---|---|
| Barriers to BILAG-2004 reliable routine use | Inaccuracy | Inefficiency | Strategy for resolution in Easy-BILAG |
| Glossary definitions for clinical items are not closely or uniformly adhered to. BILAG-2004 glossary exists as separate document to case report form and scoring document. | ● | ● | Incorporation of abbreviated glossary definitions adjacent to clinical items where possible. |
| All 97 clinical items require scoring in list format. Common clinical features must be located among many rarer manifestations. | | ● | Re-structure formatting to enhance visibility of common items in each domain. Screen for and grade rare items only when necessary. |
| Structure of scoring index does not match flow of typical clinical consultation. | | ● | Re-structure formatting to align with typical clinical consultation. |
| Deriving overall A-E domain scores requires multiple intricate algorithms which exist as separate document to the case report form | ● | ● | Uniform colour coding to simplify application of A-E scores across organ domains within single document. |
| BILAG-2004 scoring instructions (e.g. improvement criteria, trickle down rule, attribution to disease SLE activity) are not closely and uniformly adhered to. Formal training may not be universally accessible. | ● | | Supplementary key scoring instructions and self- adjudication checklist made available alongside Easy-BILAG. |

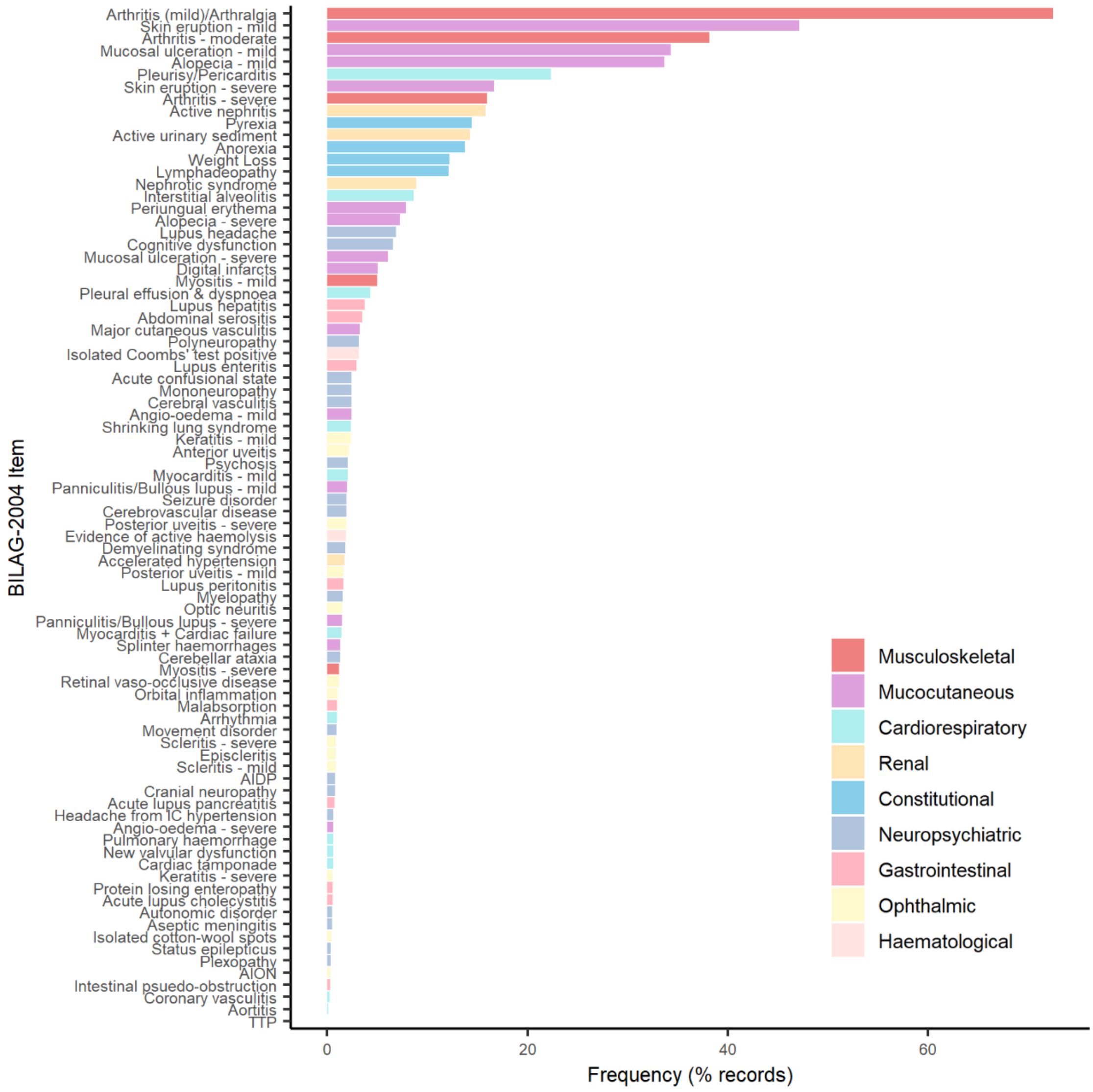


**Supplementary Figure S1: Frequency of BILAG-2004 clinical items in a biologics registry (BILAG-BR) cohort BILAG-BR.**

Histogram colour-coded by organ domain, shows frequency of all BILAG-2004 clinical items recorded as ‘improving’, ‘same’, worse’ or ‘new’ among patients with active SLE enrolled in BILAG-BR. Numerical items (full blood count indices, blood pressure, creatinine and eGFR) are mandatory to all records, irrespective of abnormality or attribution to SLE disease activity and were therefore not evaluated. Item frequency is shown as percentage of total records analysed.

| | ITEM | ABBREVIATED GLOSSARY | Nil | Imp | Same | Worse | New |
|---|---|---|---|---|---|---|---|
| Gastrointestinal | Lupus peritonitis | Serositis presenting as acute abdomen with rebound/guarding. | 0 | 1 | 2 | 3 | 4 |
| | Abdominal serositis or ascites | Not presenting as acute abdomen. | 0 | 1 | 2 | 3 | 4 |
| | Lupus enteritis / colitis | Vasculitis or inflammation of small or large bowel, with supportive imaging &/or biopsy. | 0 | 1 | 2 | 3 | 4 |
| | Malabsorption | Diarrhoea + abnormal D-xylose absorption / ↑faecal fat losses. Exclude Coeliac & gut vasculitis. | 0 | 1 | 2 | 3 | 4 |
| | Protein-losing enteropathy | See detailed glossary. | 0 | 1 | 2 | 3 | 4 |
| | Intestinal pseudo-obstruction | Subacute intestinal obstruction due to intestinal hypomotility. | 0 | 1 | 2 | 3 | 4 |
| | Lupus hepatitis | Raised transaminases, without AIH specific autoantibodies. Exclude drug- & viral hepatitis. | 0 | 1 | 2 | 3 | 4 |
| | Acute lupus cholecystitis | Exclude gallstones or infection. | 0 | 1 | 2 | 3 | 4 |
| | Acute lupus pancreatitis | Usually associated with multisystem involvement. | 0 | 1 | 2 | 3 | 4 |
| Ophthalmic | Orbital inflam / myositis / proptosis | Orbital inflammation + myositis / extra-ocular muscle swelling / proptosis. Imaging required. | 0 | 1 | 2 | 3 | 4 |
| | Keratitis - severe | Sight-threatening. Includes corneal melt and peripheral ulcerative keratitis. | 0 | 1 | 2 | 3 | 4 |
| | Keratitis - mild | Not sight-threatening. | 0 | 1 | 2 | 3 | 4 |
| | Anterior uveitis | | 0 | 1 | 2 | 3 | 4 |
| | Post. uveitis/retinal vasculitis - *severe* | Sight-threatening and/or retinal vasculitis not due to vaso-occlusive disease. | 0 | 1 | 2 | 3 | 4 |
| | Post. uveitis/retinal vasculitis - *mild* | Not sight-threatening. Not due to vaso-occlusive disease. | 0 | 1 | 2 | 3 | 4 |
| | Episcleritis | | 0 | 1 | 2 | 3 | 4 |
| | Scleritis - severe | Necrotising anterior scleritis. Ant &/or post scleritis requiring systemic therapy. | 0 | 1 | 2 | 3 | 4 |
| | Scleritis - mild | Anterior/posterior scleritis not requiring systemic steroids. | 0 | 1 | 2 | 3 | 4 |
| | Retinal / choroidal vaso-occlusive disease | See detailed glossary table. | 0 | 1 | 2 | 3 | 4 |
| | Isolated cottonwool spots | Also known as cystoid bodies. | 0 | 1 | 2 | 3 | 4 |
| | Optic neuritis | Exclude anterior ischaemic optic neuropathy. | 0 | 1 | 2 | 3 | 4 |
| | Anterior ischaemic optic neuropathy | Visual loss with pale swollen optic disc due to occlusion of posterior ciliary arteries. | 0 | 1 | 2 | 3 | 4 |

**OTHER PAGE 1 FEATURES: Record here and update the score for that domain on page one:**

| | ITEM | ABBREVIATED GLOSSARY | Nil | Imp | Same | Worse | New |
|---|---|---|---|---|---|---|---|
| Mucocutan | Angio-oedema - severe | Urticaria variant in subcut, submucosal &deep dermal tissues. Potentially life-threatening. | 0 | 1 | 2 | 3 | 4 |
| | Angio-oedema - mild | As above but not life-threatening. | 0 | 1 | 2 | 3 | 4 |
| | Panniculitis/ bullous lupus - severe | See detailed glossary table. | 0 | 1 | 2 | 3 | 4 |
| | Panniculitis/ bullous lupus - mild | Affects <9% BSA & does not fulfil any criteria for severe panniculitis. | 0 | 1 | 2 | 3 | 4 |
| | Major cutaneous vasculitis/thrombosis | Cutaneous vasculitis/thrombosis → extensive gangrene /ulceration / skin infarction. | 0 | 1 | 2 | 3 | 4 |
| | Splinter haemorrhages | | 0 | 1 | 2 | 3 | 4 |
| MSK | Myositis - *severe* | Significantly ↑ muscle enzymes with significant muscle weakness. | 0 | 1 | 2 | 3 | 4 |
| | Myositis – *mild* | Significantly ↑muscle enzymes + myalgia but no significant muscle weakness. | 0 | 1 | 2 | 3 | 4 |
| Cardiorespiratory | Myocarditis - mild | ↑ cardiac enzymes &/or ECG changes. No heart failure/arrhythmia/valve dysfunction. | 0 | 1 | 2 | 3 | 4 |
| | Myo /Endocarditis + cardiac failure | See detailed glossary table. | 0 | 1 | 2 | 3 | 4 |
| | Arrhythmias | Due to myocarditis / non-infective inflammation. ECG evidence required. | 0 | 1 | 2 | 3 | 4 |
| | New valvular dysfunction | Due to myocarditis / non-infective inflammation. Supportive imaging required. | 0 | 1 | 2 | 3 | 4 |
| | Cardiac tamponade | Supportive imaging required. | 0 | 1 | 2 | 3 | 4 |
| | Pulmonary haemorrhage/vasculitis | With haemoptysis &/or dyspnoea &/or pulmonary HTN. Imaging &/or histology required. | 0 | 1 | 2 | 3 | 4 |
| | Shrinking lung syndrome | Acute ↓ lung volumes (< 70% predicted) + normal corrected Kco. Diagphragmatic dysfunction. | 0 | 1 | 2 | 3 | 4 |
| | Aortitis | +/- dissection with supportive imaging, claudication, bruits or BP discrepancy >10 mmHg. | 0 | 1 | 2 | 3 | 4 |
| | Coronary vasculitis | Imaging evidence of non-atheromatous coronary narrowing/ obstruction /aneurysm. | 0 | 1 | 2 | 3 | 4 |
| Neuropsychiatric | Aseptic meningitis | See detailed glossary table. | 0 | 1 | 2 | 3 | 4 |
| | Cerebral vasculitis | With features of vasculitis in another system. Supportive imaging &/or biopsy required. | 0 | 1 | 2 | 3 | 4 |
| | Demyelinating syndrome | Discrete white matter lesion + neurological deficit. Ideally ≥1 prior recorded event. Exclude MS. | 0 | 1 | 2 | 3 | 4 |
| | Myelopathy | Acute onset, rapidly evolving paraparesis, quadriparesis and/or sensory level. Exclude SOL. | 0 | 1 | 2 | 3 | 4 |
| | Acute confusional state | See detailed glossary table. | 0 | 1 | 2 | 3 | 4 |
| | Psychosis | Delusions &/or hallucinations. Excludes primary psychotic disorder, drugs or during delirium. | 0 | 1 | 2 | 3 | 4 |
| | Acute inflammatory demyelinating polyradiculoneuropathy | See detailed glossary table. | 0 | 1 | 2 | 3 | 4 |
| | Cranial neuropathy | Exclude optic neuropathy which is classified under ophthalmic system. | 0 | 1 | 2 | 3 | 4 |
| | Plexopathy | Supportive electrophysiology study required. | 0 | 1 | 2 | 3 | 4 |
| | Cognitive dysfunction | Sufficient to impair ADLs. Includes attention, memory, language, visuospatial, psychomotor. | 0 | 1 | 2 | 3 | 4 |
| | Status epilepticus | A seizure or seizures lasting >30 minutes without full recovery to baseline. | 0 | 1 | 2 | 3 | 4 |
| | Cerebrovascular disease | Not vasculitis. See detailed glossary table. | 0 | 1 | 2 | 3 | 4 |
| | Movement disorder | Exclude drug-induced. | 0 | 1 | 2 | 3 | 4 |
| | Autonomic disorder | See detailed glossary table. | 0 | 1 | 2 | 3 | 4 |
| | Cerebellar ataxia | Cerebellar ataxia in isolation of other CNS features. Usually subacute presentation. | 0 | 1 | 2 | 3 | 4 |
| | Severe lupus headache | Unremitting, Disabling, unresponsive to narcotics, >3 days. Exclude SOL and CNS infection. | 0 | 1 | 2 | 3 | 4 |
| | Headache from intracranial hypertension | Exclude cerebral sinus thrombosis. | 0 | 1 | 2 | 3 | 4 |
| Haem | TTP | Micro-angiopathic haemolytic anaemia + thrombocytopenia. Other causes excluded. | 0 | 1 | 2 | 3 | 4 |
| | Coombes positive (isolated) | Without evidence of haemolysis. | Negative | | Positive | | |

© University of Leeds

**Supplementary Figure S2: Easy-BILAG page 2 captures infrequent and rare BILAG-2004 clinical items.**

Easy-BILAG page 2 in full demonstrates structure of the scoring template for uncommon and rare SLE features not captured by page 1. Items are organised within tables by organ domain. Gastrointestinal and Ophthalmic domains are represented in full, followed by infrequently scored items from the Mucocutaneous (Mucocutan), Musculoskeletal (MSK), Cardiorespiratory, Neuropsychiatric and Haematological (Haem) domains, signposted on page 1. Scoring of items from ‘not present’ (0) to ‘new’ (4) is assisted by colour coding which translates to overall organ domain scores from A (blue), B (pink), C (yellow) to D or E (white) as directed by the key on Easy-BILAG page 1 (See Figure 1). If any page 2 items are recorded, an updated domain score for each organ domain is assigned in the free text space provided on Easy-BILAG page 1.

### EASY -BILAG

Self- Adjudication Checklist

1. All items scored are attributable to active SLE

2. All abnormal blood and urine results scored are attributable to active SLE

3. The ‘trickle down rule’ has been applied:
   Any item scored as ‘severe’ should also be scored as ‘mild’

4. If completed at monthly intervals confirm:
   Any item scored ‘same’ or ‘worse’ was also scored last month
   Any item scored ‘new’ last month is also scored this month

5. Items scored as improving have been *either* consistently improving over 2 weeks *or* fully resolved for at least 7 days *and* improvement is sufficient to consider alteration in therapy

**Supplementary Figure S3. Optional Easy-BILAG self-adjudication check-list.**
An optional five-item checklist to follow completion of Easy-BILAG is designed to assist use of Easy-BILAG in clinical research and mitigate against common inconsistencies which arise in BILAG-2004 assessments during clinical trials.

**Supplementary table S2. Characteristics of professionals participating in Easy-BILAG validation exercise**

| | All<br><br>n = 33 | Standard BILAG-2004 Index<br>n = 17 | Easy BILAG<br><br>n = 16 |
|---|---|---|---|
| **Clinical Role** | | | |
| *Consultant Rheumatologist* | 12 (36.4) | 5 (29.4) | 7 (43.8) |
| *Speciality Trainee* | 15 (45.5) | 9 (52.9) | 6 (37.5) |
| *Clinical Academic* | 3 (9.1) | 1 (5.9) | 2 (12.5) |
| *Clinical Nurse Specialist* | 3 (9.1) | 2 (11.8) | 1 (6.3) |
| **Prior BILAG-2004 training** | 18 (54.5) | 8 (47.1%) | 10 (62.5) |
| **Workplace** | | | |
| *General hospital* | 13 (39.4) | 8 (47.1) | 5 (31.3) |
| *Tertiary centre* | 20 (60.6) | 9 (52.9) | 11 (68.8) |
| **Current BILAG-2004 use** | | | |
| *Regularly* | 10 (27.3) | 3 (17.6) | 7 (43.8) |
| *Occasional or rarely* | 23 (69.7) | 14 (82.4) | 9 (56.3) |

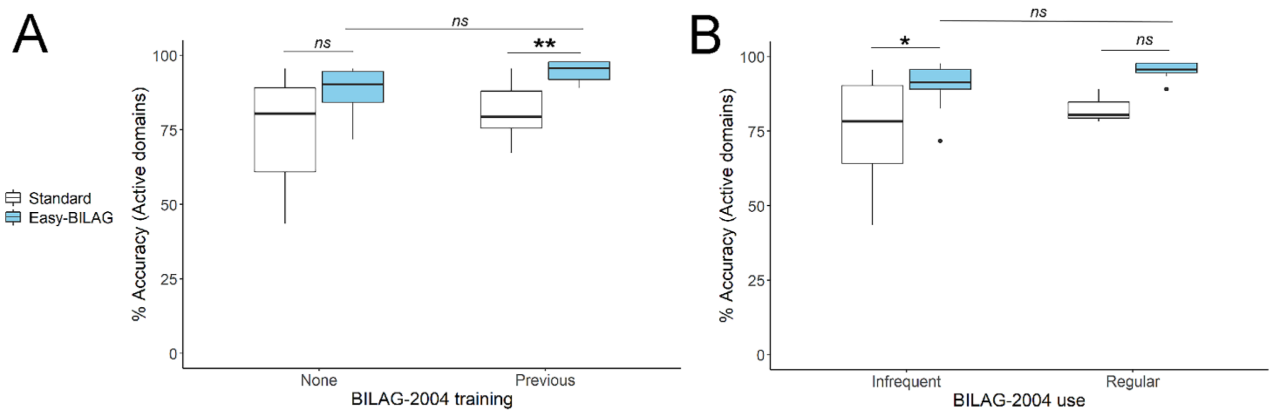


**Supplementary Figure S4. Accuracy of Easy-BILAG by prior BILAG-2004 training and experience.**

Boxplots show performance of Easy-BILAG (blue) versus standard format BILAG-2004 (white) across different levels of prior BILAG-2004 training and experience 2004 in the self-timed validation workbook exercise. For clinicians with prior BILAG-2004 training, scoring accuracy (% accuracy; *median* ($Q_1$, $Q_2$)) against model answers for active organ domains (A) was significantly higher using Easy-BILAG (n = 10; 95.7 (91.3, 97.8)) than standard format BILAG-2004 (n = 8; 79.3 (75.0, 89.1), p = 0.01). Among those without prior training there was also a lesser effect (p = 0.2) but accuracy of clinicians using Easy-BILAG was not significantly different between those with and without prior training (p = 0.30). Scoring accuracy for active domains among clinicians who currently use BILAG-2004 infrequently in practice (B) was significantly higher with Easy-BILAG (n = 9; 91.3 (89.1, 95.7)) than with standard format BILAG-2004 (n = 15; 78.3 (62.5, 89.1), p = 0.05). Clinicians using BILAG-2004 regularly also showed trend to higher accuracy with Easy-BILAG (n = 7; 95.6 (94.6, 97.8)) and standard format BILAG-2004 (n = 2; median = 84.8; p = 0.09) though numbers for comparison were smaller. Scoring accuracy using Easy-BILAG was not significantly different between clinicians using existing BILAG-2004 regularly and infrequently in current practice (p = 0.15). ** p ≤ 0.01 * p ≤ 0.05, ns – non significant, p > 0.05
